# The Impact of Myosteatosis on Cardiac Function in a Healthy Population: Insights from Abdominal CT Imaging

**DOI:** 10.1101/2024.08.02.24311439

**Authors:** Myung Jin Kim, Hyo-Jung Nam, Yun Kyung Cho, Eun Hee Kim, Min Jung Lee, Woo Je Lee, Hong-Kyu Kim, Chang Hee Jung

**Author notes:** Corresponding Authors 1) Chang Hee Jung, MD, PhD (https://orcid.org/0000-0003-4043-2396) Department of Internal Medicine, Asan Medical Center, University of Ulsan College of Medicine, 88, Olympic-ro 43-gil, Songpa-gu, Seoul, 05505, Republic of Korea.) Hong-Kyu Kim, MD, PhD (https://orcid.org/0000-0002-7606-3521) Health Screening and Promotion Center, Asan Medical Center, University of Ulsan College of Medicine, 88, Olympic-ro 43-gil, Songpa-gu, Seoul, 05505, Republic of Korea.

## Abstract

**Background:** Ectopic fat deposition in skeletal muscle, termed myosteatosis, is a key factor in insulin resistance and contributes to various metabolic disturbances. This study evaluated the association between myosteatosis, assessed via abdominal computed tomography, and cardiac function in an asymptomatic Korean population.

**Methods:** This cross-sectional study included 7,716 participants (4,902 [63.5%] men and 2,814 [36.5%] women, mean age 53.2 ± 8.0) who underwent routine health check-ups at Asan Medical Center (Seoul, Korea). Myosteatosis was evaluated by segmenting the total abdominal muscle area (TAMA) at the L3 vertebral level into normal-attenuation muscle area (NAMA), low-attenuation muscle area (LAMA), and inter-/intra-muscular adipose tissue (IMAT). Patients were categorized into quartiles based on the NAMA/TAMA index, calculated by dividing the NAMA by TAMA and multiplying by 100. Cardiac function was assessed by transthoracic echocardiography.

**Results:** Higher NAMA/TAMA index levels were associated with decreased absolute values of the E/E’ ratio and increased E/A ratio in both men and women. Multiple linear regression analysis revealed a significant correlation between the NAMA/TAMA index and both the E/A and E/E’ ratios after adjusting for covariates. No significant correlation was found with left ventricular (LV) ejection fraction or LV mass index.

**Conclusions:** The degree of myosteatosis was significantly associated with diastolic function in an asymptomatic population, while systolic function remained unaffected.

## 1. Introduction

Ectopic fat infiltration in non-adipose organs is now recognized as an important contributor to the pathogenesis of insulin resistance and metabolic dysregulations. ^1,2^ Specifically, ectopic fat deposition in skeletal muscle, termed myosteatosis, may deteriorate muscle quality and induce systemic insulin resistance, considering that skeletal muscle functions as the primary organ for insulin action and glucose metabolism. ^1^ Our previous study demonstrated that myosteatosis was independently associated with systemic and hepatic insulin resistance, regardless of body size and the presence of other metabolic risk factors. ^3^

Computed tomography (CT) is commonly used to evaluate myosteatosis as a non-invasive alternative to muscle biopsy, providing reliable results that correlate with biopsy findings. ^4,5^ Given that CT density decreases with fatty tissue infiltration, the total abdominal muscular area (TAMA) in the CT axial image can be separated into three components based on CT density: normal-attenuation muscle area (NAMA), low-attenuation muscle area (LAMA), and inter- or intramuscular adipose tissue (IMAT). ^6^ The NAMA/TAMA index, which has been proposed as a reliable measure of muscle quality, is calculated by dividing NAMA by TAMA and multiplying by 100, indicating the proportion of healthy muscle with minimal fat infiltration. ^5^ Individuals with a higher NAMA/TAMA index were shown to have a lower risk of subclinical coronary atherosclerosis^7^ and more favorable metabolic characteristics, including less degree of systemic insulin resistance. ^3^

Insulin-resistant conditions can induce cardiac dysfunction similar to that of diabetic cardiomyopathy, even in individuals without clinical diabetes mellitus. ^8^ A significant association between the left ventricular (LV) diastolic function and the homeostasis model assessment of insulin resistance (HOMA-IR) has been consistently demonstrated in individuals with or without diabetes. ^9–13^ These findings suggest that insulin resistance independently predisposes individuals to the development of cardiac dysfunction.

Given that myosteatosis is an independent factor inducing insulin resistance, we hypothesized that a relationship might exist between myosteatosis and cardiac function; however, this association has not yet been studied. Our study was designed to investigate the impact of myosteatosis, assessed by abdominal CT, on cardiac function in a healthy asymptomatic population.

## 2. Methods

### 2.1. Study population

This cross-sectional study included 9,519 individuals who underwent both abdominal CT and transthoracic echocardiography as part of routine health checkups at the Asan Medical Center (Seoul, Republic of Korea) between January 2012 and December 2013. Participants were excluded if they had any of the conditions listed below: history of malignancy; history of cardiovascular disease; history of cerebrovascular accidents; history of lung disease; chronic kidney disease; liver cirrhosis; overt or subclinical thyroid dysfunction; or current glucocorticoid or sex hormone replacement. Detailed exclusion criteria are described in the **Supplementary Methods**.

### 2.2. Laboratory and anthropometric assessments

Sociodemographic and lifestyle information was collected based on standardized questionnaires. Drinking habits were defined by the frequency of alcohol consumption, with individuals drinking ≥2 days/week classified as moderate drinkers. Smoking status was identified as either non-current or current. Regular exercise was defined as performing 30 minutes of moderate-intensity aerobic exercise for ≥5 days/week, 20 minutes of vigorous-intensity exercise for ≥3 days/week, or resistance exercise for ≥3 days/week. Body mass index (BMI) was calculated by dividing weight (kg) by height (m) squared. Direct segmental multi-frequency bioelectrical impedance analysis (InBody 720, InBody CO., Ltd., Seoul, Korea) was used to measure body composition. Hypertension was defined as having systolic/diastolic blood pressure (BP) above 140/90 mmHg or ongoing use of antihypertensive drugs. Diabetes was defined as one of the following: fasting plasma glucose (FPG) ≥7.0 mmol/L (≥ 126 mg/dL); glycated hemoglobin (HbA1c) ≥ 6.5%; or the use of anti-diabetic medication. More detailed explanations of the anthropometric and laboratory assessments are provided in the **Supplementary Methods**.

### 2.3. Evaluation of myosteatosis

Myosteatosis was assessed using an axial image of abdominal CT scan at the L3 vertebrae level. An artificial intelligence program was employed to automatically analyze body composition from CT images. ^14^ This automated system segmented the body part on axial images into the subcutaneous fat, visceral fat, and TAMA. TAMA included all visible muscles on the L3 axial image. **Figure S1** graphically describes how body composition in the axial CT image was analyzed.

TAMA was further divided into three segments based on mean CT densities (Hounsfield Unit [HU]): (1) NAMA ranged from +30 to +150 HU, indicating minimal fat content; (2) LAMA ranged from −29 to +29 HU, representing muscle with a higher fat content; and (3) IMAT ranged from −190 to −30 HU. Skeletal muscle area (SMA) was defined as the combined area of NAMA and LAMA, spanning from −29 to +150 HU. As a myosteatosis index, we used the NAMA/TAMA index, which was calculated by dividing NAMA by TAMA and multiplying the result by 100. ^3^ A higher NAMA/TAMA index value represents a higher degree of good-quality muscle. ^15^

### 2.4. Evaluation of cardiac function

Transthoracic echocardiographic evaluation was performed by experienced sonographers or three experienced cardiologists using commercially available ultrasound machines (iE33, EPIC; Philips Medical Systems, Andover, MA). Conventional two-dimensional and Doppler images were obtained following the guidelines of the American Society of Echocardiography (ASE) and the European Association of Cardiovascular Imaging. ^16^ End-systolic internal diameter (LVIDs), end-diastolic internal diameter (LVIDd), interventricular septal thickness at diastole (IVSd), posterior wall thickness at diastole of the LV (LVPWd), and left atrial diameter were measured from the parasternal long-axis view guided by 2D M-mode acquisitions. The end-systolic volume, end-diastolic volume, and the LV ejection fraction (LVEF) were evaluated using the biplane Simpson method. ^16^ LV mass index was calculated from the anatomically verified cube formula using the values measured by M-mode. ^16^ Pulsed-wave Doppler images were captured at the mitral valve tips from apical four-chamber 2D views to measure flow velocities during the peak early diastolic (E-wave) and peak late diastolic (A-wave) phases, allowing for the calculation of the E/A ratio as an indicator of diastolic filling. Tissue Doppler Imaging (TDI) was performed to estimate the early peak velocity (E’) of the septal mitral annulus. According to ASE Recommendations for the Evaluation of Left Ventricular Diastolic Function, ^17^ diastolic function was assessed based on the E wave, E/A ratio, E’ and E/E’ ratio.

### 2.5. Statistical analysis

All statistical analyses were conducted independently for men and women due to variations in muscle characteristics. ^18^ Based on the quartiles of the NAMA/TAMA index, the participants were categorized into four groups. Baseline clinical and echocardiographic data for each quartile group were presented as mean ± standard deviation (SD) for continuous variables and number and percentage for categorical variables. Differences between groups were determined using analysis of variance (ANOVA) with Scheffe’s method for post-hoc analysis of continuous variables and the Kruskal–Wallis test with the Dunn procedure for those with skewed distributions. A chi-square test was used to assess categorical variables.

Multiple linear regression analysis was conducted to assess the association between the NAMA/TAMA index and key indicators of cardiac function, including LVEF, LV mass index, E/A ratio, and E/E’ ratio. The analyses were performed with four sequential adjustment models, each incorporating different sets of adjustment variables: the unadjusted model; model 1, which adjusted for age; model 2, which adjusted for smoking status, drinking habits, regular exercise, hypertension, and diabetes mellitus in conjunction with factors included in model 1; and model 3, which adjusted for gamma-glutamyl transferase (GGT), triglyceride, low-density lipoprotein cholesterol (LDL-C), high-sensitivity C-reactive protein (hsCRP), and visceral-subcutaneous fat ratio (VSR) in addition to the variables of model 2. Pearson’s correlation coefficients were used to evaluate the correlation between the NAMA/TAMA index and echocardiographic indices.

All statistical analyses were performed using SPSS software version 21.0 for Windows (IBM, Inc., Armonk, NY, USA). P-values below 0.05 were regarded as statistically significant.

### 2.6. Ethics statement

The study protocol was approved by the Institutional Review Board of Asan Medical Center (IRB No. 2023-1360). Given the retrospective nature of the study utilizing pre-existing, de-identified clinical data, the requirement for written informed consent was waived. The study was conducted following the ethical standards of the 1964 Declaration of Helsinki and its later amendments.

## 3. Results

### 3.1. Baseline characteristics of the study population

A total of 7,716 participants (4,902 [63.5%] men and 2,814 [36.5%] women) were included in the final analysis (**Fig. 1**). The cutoff values of the NAMA/TAMA index quartiles were 71.93, 77.38, and 81.53 in males and 65.02, 71.79, and 77.18 in females. The mean age of the participants was 53.2±8.0 years, and the mean BMI was 24.1±3.0 kg/m^2^.

**Figure 1.**
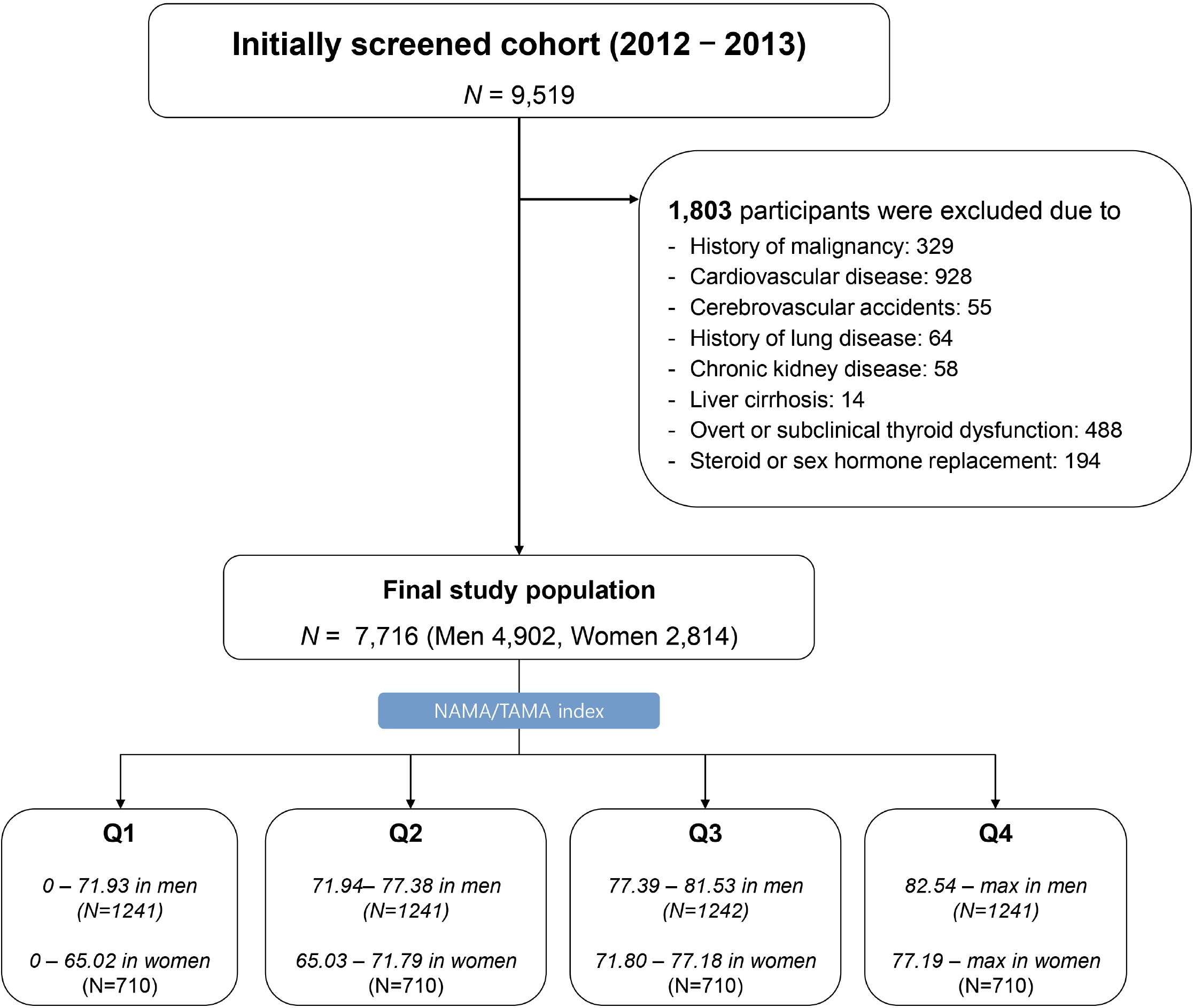
Study population. NAMA, normal-attenuation muscle area; TAMA, total abdominal muscle area.

The baseline characteristics and body measurements according to the NAMA/TAMA index quartile groups are presented in **Table 1**. Overall, patients with higher NAMA/TAMA index values exhibited more favorable metabolic profiles, such as lower BP, better glycemic and lipid profiles, and lower HOMA-IR. Regarding body composition, percent body fat and VSR decreased with higher NAMA/TAMA index levels. The mean values of TAMA, LAMA, LAMA/BMI, and IMAT showed downward trends with increased NAMA/TAMA index, while SMA, SMA/BMI, NAMA, and NAMA/BMI showed upward trends.

**Table 1.** Baseline clinical and biochemical characteristics of all participants according to the quartiles of NAMA/TAMA index.

|  | Men (N = 4,902) |  |  |  |  | Women (N = 2,814) |  |  |  |  |
| --- | --- | --- | --- | --- | --- | --- | --- | --- | --- | --- |
|  | Q1 | Q2 | Q3 | Q4 | P | Q1 | Q2 | Q3 | Q4 | P |
| N | 1225 | 1226 | 1226 | 1225 |  | 703 | 704 | 704 | 703 |  |
| Age (years) | 55.7±8.5 | 53.8±7.8 | 52.4±7.1 | 50.2±7.3 | <0.001 | 58.1±8.2 | 54.1±7.5 | 52.6±7.0 | 49.0±6.8 | <0.001 |
| Body mass index (kg/m<sup>2</sup>) | 26.0±3.2 | 24.9±2.5 | 24.5±2.4 | 23.6±2.5 | <0.001 | 24.7±3.3 | 23.2±2.7 | 22.4±2.6 | 21.0±2.3 | <0.001 |
| Systolic BP (mmHg) | 125.5±11.9 | 124.1±11.9<sup>a</sup> | 123.4±11.5<sup>ab</sup> | 122.2±11.6<sup>b</sup> | <0.001 | 122.2±14.0 | 117.3±13.6 | 115.3±13.4 | 113.1±13.0 | <0.001 |
| Diastolic BP (mmHg) | 80.8±9.2<sup>a</sup> | 80.2±9.2<sup>a</sup> | 79.8±9.1<sup>ab</sup> | 78.8±9.3<sup>b</sup> | <0.001 | 75.3±9.7 | 73.2±9.6<sup>a</sup> | 72.4±16.2<sup>a</sup> | 71.0±10.3 | <0.001 |
| Current smoker (%) | 383 (31.4) | 407 (33.2) | 416 (34.0) | 429 (35.0) | 0.049 | 22 (3.1) | 22 (3.1) | 19 (2.7) | 22 (3.1) | 0.879 |
| Moderate drinker (%) | 608 (49.8) | 646 (52.7) | 599 (48.9) | 537 (43.9) | 0.001 | 43 (6.1) | 42 (6.0) | 30 (4.3) | 42 (6.0) | 0.354 |
| Hypertension (%) | 448 (36.6) | 353 (28.8) | 331 (27.0) | 231 (18.9) | <0.001 | 191 (27.2) | 119 (16.9) | 110 (15.6) | 55 (7.8) | <0.001 |
| Diabetes mellitus (%) | 169 (13.8) | 155 (12.6) | 101 (8.2) | 99 (8.1) | <0.001 | 48 (6.8) | 35 (5.0) | 25 (3.6) | 16 (2.3) | <0.001 |
| Dyslipidemia therapy (%) | 192 (15.7) | 167 (13.6) | 138 (11.3) | 105 (8.6) | <0.001 | 114 (16.2) | 87 (12.4) | 67 (9.5) | 41 (5.8) | <0.001 |
| FPG (mg/dL) | 107.7±23.8<sup>a</sup> | 105.7±22.2<sup>a</sup> | 102.6±17.6<sup>b</sup> | 101.8±18.0<sup>b</sup> | <0.001 | 101.1±18.1 | 97.9±14.9<sup>a</sup> | 96.9±14.6<sup>a</sup> | 94.2±11.6 | <0.001 |
| HbA1c (%) | 5.82±0.88 | 5.74±0.77 | 5.63±0.61<sup>a</sup> | 5.60±0.63<sup>a</sup> | <0.001 | 5.72±0.68 | 5.61±0.60<sup>a</sup> | 5.58±0.55<sup>a</sup> | 5.40±0.42 | <0.001 |
| Total cholesterol (mg/dL) | 191.3±34.8<sup>a</sup> | 194.3±34.6<sup>a</sup> | 194.0±33.9<sup>ab</sup> | 195.6±34.1<sup>b</sup> | 0.018 | 204.3±36.9<sup>a</sup> | 201.0±36.1<sup>a</sup> | 199.3±33.4<sup>a</sup> | 193.6±32.5 | <0.001 |
| Triglyceride (mg/dL) | 125 (90–169) | 123 (89–171) | 118 (86–162) | 114 (81–157) | <0.001 | 98 (73–133) | 90 (67–132) | 87 (65–126) | 76 (56–105) | <0.001 |
| LDL-C (mg/dL) | 121.6±31.4 | 123.5±30.1 | 124.0±30.3 | 124.6±30.1 | 0.083 | 129.8±32.9<sup>a</sup> | 125.5±32.2<sup>ab</sup> | 124.3±29.4<sup>b</sup> | 117.0±29.3 | <0.001 |
| HDL-C (mg/dL) | 50.1±11.8<sup>a</sup> | 50.5±12.0<sup>a</sup> | 51.0±13.0<sup>a</sup> | 52.8±13.6 | <0.001 | 58.1±13.4 | 60.5±15.1<sup>a</sup> | 60.9±14.9<sup>a</sup> | 64.4±15.3 | <0.001 |
| GGT (U/L) | 47.0±69.4<sup>a</sup> | 43.9±45.2<sup>ab</sup> | 40.9±41.8<sup>bc</sup> | 35.8±35.9<sup>c</sup> | <0.001 | 22.9±39.8<sup>a</sup> | 19.7±18.3<sup>ab</sup> | 18.9±17.2<sup>b</sup> | 14.8±9.8 | <0.001 |
| hsCRP (mg/L) | 0.08 (0.05–0.17) | 0.07 (0.04–0.14) | 0.07 (0.04–0.13) | 0.06 (0.04–0.11) | <0.001 | 0.07 (0.04–0.15) | 0.06 (0.04–0.12) | 0.06 (0.04–0.10) | 0.05 (0.03–0.11) | <0.001 |
| HOMA-IR | 1.56 (0.92–2.33) | 1.37 (0.80–2.05) | 1.28 (0.76–1.93) | 1.15 (0.64–1.79) | <0.001 | 1.33 (0.77–1.97) | 1.12 (0.69–1.67) | 0.98 (0.58–1.57) | 0.84 (0.52–1.35) | <0.001 |
| **Body composition** |  |  |  |  |  |  |  |  |  |  |
| Percent body fat (%) | 25.2±4.9 | 22.5±4.3 | 21.2±4.1 | 18.7±4.4 | <0.001 | 34.1±5.6 | 30.5±5.2 | 28.3±5.3 | 24.8±5.4 | <0.001 |
| VSR | 1.27±0.53<sup>a</sup> | 1.23±0.52<sup>a</sup> | 1.16±0.47 | 1.02±0.48 | <0.001 | 0.58±0.27 | 0.50±0.27 | 0.46±0.24 | 0.37±0.23 | <0.001 |
| TAMA, cm<sup>2</sup> | 175.4±24.2 | 174.5±22.4 | 174.1±21.1 | 173.6±21.4 | 0.256 | 120.5±15.1 | 117.1±13.8<sup>a</sup> | 115.9±13.3<sup>ab</sup> | 114.4±13.2<sup>b</sup> | <0.001 |
| SMA, cm<sup>2</sup> | 161.9±23.1 | 165.2±21.5<sup>a</sup> | 166.8±20.4<sup>ab</sup> | 168.5±20.8<sup>b</sup> | <0.001 | 106.1±13.3<sup>a</sup> | 108.0±13.0<sup>ab</sup> | 109.2±12.7<sup>bc</sup> | 109.9±13.5<sup>c</sup> | <0.001 |
| SMA/BMI | 6.24±0.66 | 6.63±0.61 | 6.81±0.58 | 7.15±0.65 | <0.001 | 4.32±0.52 | 4.68±0.51 | 4.90±0.51 | 5.26±0.62 | <0.001 |
| NAMA, cm<sup>2</sup> | 116.1±18.5 | 130.6±17.0 | 138.3±16.7 | 146.4±18.4 | <0.001 | 68.8±10.8 | 80.4±9.7 | 86.5±10.8 | 92.6±11.2 | <0.001 |
| NAMA/BMI | 4.48±0.63 | 5.24±0.50 | 5.64±0.49 | 6.21±0.61 | <0.001 | 2.81±0.49 | 3.49±0.39 | 3.88±0.41 | 4.43±0.54 | <0.001 |
| LAMA, cm<sup>2</sup> | 45.8±10.4 | 34.6±5.5 | 28.6±4.4 | 22.1±4.1 | <0.001 | 37.3±8.3 | 27.6±4.3 | 22.8±3.3 | 17.6±3.2 | <0.001 |
| LAMA/BMI | 1.76±0.30 | 1.39±0.17 | 1.16±0.14 | 0.93±0.14 | <0.001 | 1.51±0.26 | 1.19±0.16 | 1.02±0.13 | 0.84±0.13 | <0.001 |
| IMAT, cm<sup>2</sup> | 13.5±5.5 | 9.3±2.7 | 7.3±2.1 | 5.1±1.8 | <0.001 | 14.4±5.2 | 9.1±2.4 | 6.7±1.7 | 4.3±1.5 | <0.001 |
| NAMA/TAMA index | 66.2±5.6 | 74.9±1.5 | 79.4±1.2 | 84.3±2.1 | <0.001 | 57.3±6.8 | 68.7±2.0 | 74.6±1.5 | 80.9±2.6 | <0.001 |

Values were presented as mean ± standard deviation and n (%) for categorical variables, where appropriate.
<sup>a,b,c</sup>The same superscripts imply a statistically insignificant difference between those values in post hoc analysis. Otherwise, post hoc analysis revealed significant differences between each group.
**Abbreviations:** BP, blood pressure; FPG, fasting plasma glucose; HbA1c, hemoglobin A1c; LDL-C, low-density lipoprotein-cholesterol; HDL-C, high-density lipoprotein-cholesterol; GGT, gamma-glutamyltransferase; hsCRP, high-sensitivity C-reactive protein; HOMA-IR, Homeostatic Model Assessment for Insulin Resistance; VSR, visceral to subcutaneous fat ratio; TAMA, total abdominal muscle area; NAMA, normal-attenuation muscle area; LAMA, low-attenuation muscle area; IMAT, intermuscular adipose tissue.

### 3.2. Echocardiographic findings according to the quartiles of NAMA/TAMA index

Echocardiographic characteristics are presented in **Table 2**. The LVPWd and IVSd were significantly higher in participants with lower NAMA/TAMA index values in both men and women. There were significant differences in the LV mass index between the NAMA/TAMA index groups in women, but not in men. While LVEF showed no significant differences among men, it demonstrated a significant decreasing trend with higher NAMA/TAMA index levels in women. Significant differences were observed in left atrial (LA) diameter, E wave, A wave, E/A ratio, and E/E’ ratio across the NAMA/TAMA index quartiles (all p <0.001). Specifically, as the NAMA/TAMA index decreased, indicating a greater degree of myosteatosis, the E wave and E/A ratio significantly decreased, whereas the LA diameter, A wave, and E/E’ ratio increased, demonstrating diastolic dysfunction.

**Table 2.** Echocardiographic variables of the study population according to the quartiles of NAMA/TAMA index.

|  | Men |  |  |  |  | Women |  |  |  |  |
| --- | --- | --- | --- | --- | --- | --- | --- | --- | --- | --- |
|  | Q1 | Q2 | Q3 | Q4 | P | Q1 | Q2 | Q3 | Q4 | P |
| Echocardiographic findings |  |  |  | 0 |  |  |  |  |  |  |
| LVIDd (mm) | 49.4±4.1 | 49.4±3.9 | 49.2±3.7 | 49.1±3.7 | 0.061 | 46.7±3.9<sup>a</sup> | 46.2±3.7<sup>ab</sup> | 46.4±3.4<sup>ab</sup> | 45.8±3.5<sup>b</sup> | <0.001 |
| LVPWd (mm) | 9.13±1.06<sup>a</sup> | 9.04±1.02<sup>ab</sup> | 8.96±1.26<sup>bc</sup> | 8.85±1.01<sup>c</sup> | <0.001 | 8.23±1.05 | 8.02±1.20 | 7.87±1.08 | 7.58±1.00 | <0.001 |
| IVSd (mm) | 9.21±1.16 | 9.02±1.19<sup>a</sup> | 8.92±1.12<sup>ab</sup> | 8.81±1.16<sup>b</sup> | <0.001 | 8.28±1.23 | 7.92±1.23<sup>a</sup> | 7.80±1.20<sup>a</sup> | 7.46±1.12 | <0.001 |
| LA diameter (mm) | 37.4±4.2 | 36.6±4.0 | 35.9±4.1 | 35.0±4.2 | <0.001 | 35.1±4.0 | 33.7±3.8 | 32.9±3.7 | 31.8±3.8 | <0.001 |
| LVEF (%) | 62.7±4.0 | 63.0±4.0 | 62.9±3.9 | 62.6±3.5 | 0.109 | 64.1±3.9 | 64.3±3.8 | 64.0±3.9 | 63.8±3.8 | 0.046 |
| LV mass index (g/m<sup>2</sup>) | 85.4±16.1 | 85.5±16.3 | 84.8±15.5 | 84.1±15.7 | 0.162 | 78.9±16.3 | 75.6±16.4<sup>a</sup> | 75.1±16.2<sup>a</sup> | 71.4±15.2 | <0.001 |
| MV pulsed doppler |  |  |  |  |  |  |  |  |  |  |
| E wave (cm/sec) | 58.6±13.1<sup>a</sup> | 59.2±12.8<sup>ab</sup> | 60.3±13.0<sup>bc</sup> | 60.9±12.7<sup>c</sup> | <0.001 | 65.5±14.8<sup>a</sup> | 67.4±14.1<sup>ab</sup> | 69.0±13.8<sup>b</sup> | 70.9±13.9 | <0.001 |
| A wave (cm/sec) | 61.3±13.4 | 58.8±12.2 | 57.3±12.6 | 54.1±12.0 | <0.001 | 66.6±14.9 | 62.4±14.3 | 59.5±15.1 | 55.0±13.9 | <0.001 |
| E/A ratio | 0.99±0.28 | 1.04±0.29 | 1.09±0.30 | 1.17±0.33 | <0.001 | 1.02±0.30 | 1.13±0.33 | 1.22±0.37 | 1.36±0.41 | <0.001 |
| MV DTI parameter |  |  |  |  |  |  |  |  |  |  |
| E/E' ratio | 9.46±2.50 | 9.06±2.21<sup>a</sup> | 8.93±2.18<sup>a</sup> | 8.54±2.06 | <0.001 | 10.70±2.89 | 10.01±2.47 | 9.60±2.36 | 8.80±2.18 | <0.001 |

Values were presented as mean ± standard deviation.
<sup>a,b,c</sup>The same superscripts imply a statistically insignificant difference between those values in post hoc analysis. Otherwise, post hoc analysis revealed significant differences between each group.
**Abbreviations:** LVIDd, left ventricular internal diameter at end-diastole; LVPWd, left ventricular posterobasal free wall thickness at end-diastole; IVSd, interventricular septum thickness at end-diastole; LA, left atrium; LVEF, left ventricular ejection fraction; LV, left ventricle; MV, mitral valve; DTI, doppler tissue imaging.

### 3.3. Association between NAMA/TAMA index and echocardiographic variables after adjustment

Multiple regression analysis revealed that the NAMA/TAMA index was positively associated with the E/A ratio and negatively associated with the E/E’ ratio in both unadjusted and adjusted models (**Table 3**). After adjusting for covariates, these associations remained significant for both the E/A ratio (β = 0.003, p <0.001 in men and β = 0.002, p =0.001 in women in model 3) and E/E’ ratio (β = −0.012, p =0.015 in men and β = −0.026, p <0.015 in the model 3). Significant associations were observed between the NAMA/TAMA index and the LVEF (p = 0.017) or LV mass index (p < 0.001) in women, but they were not maintained after adjusting (Model 3) for covariates. The scatter plot depicting the relationship between the NAMA/TAMA index and the E/E’ ratio and E/A ratio is shown in **Fig. 2**.

**Figure 2.**
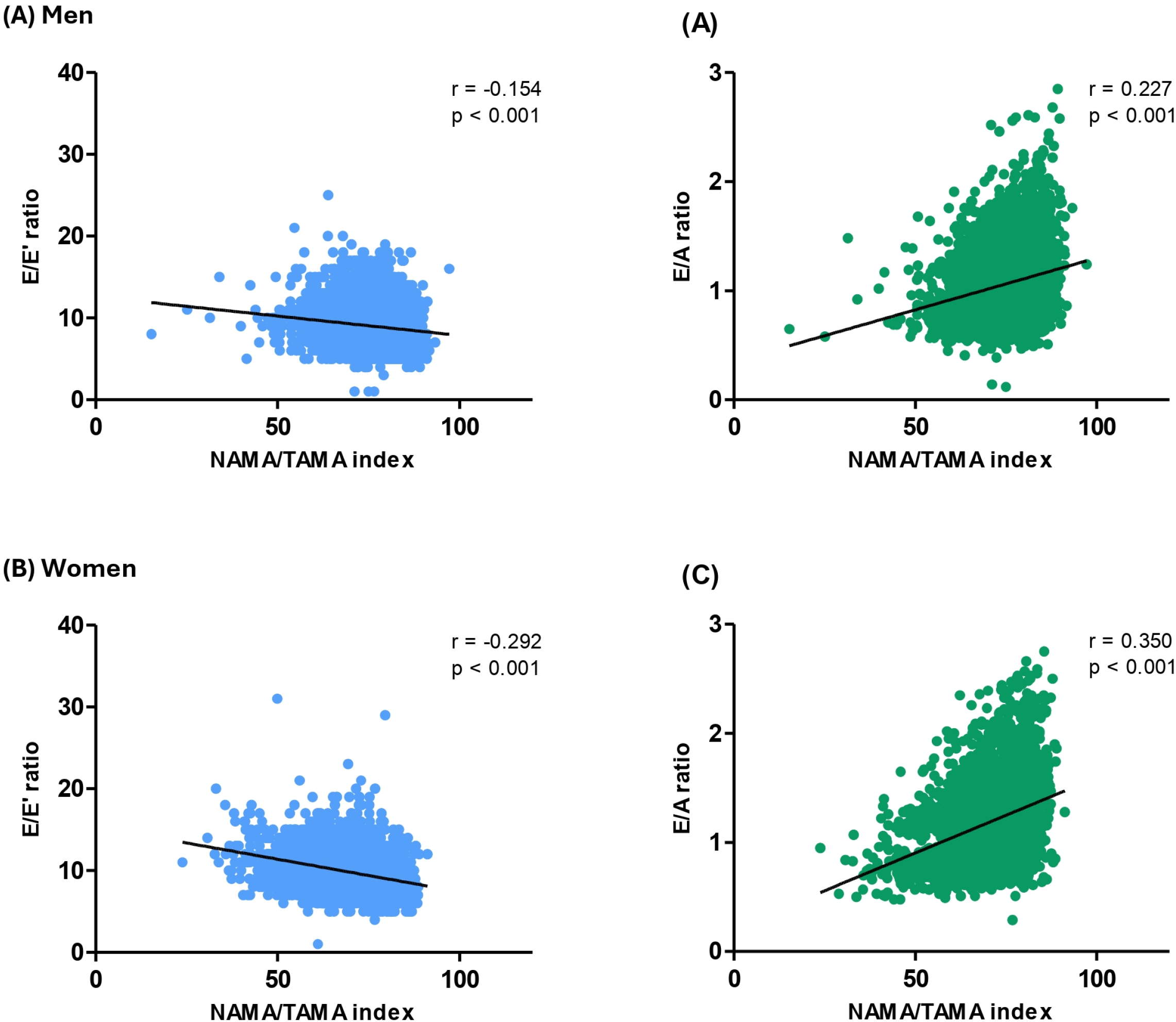
Scatter plot of the correlation between NAMA/TAMA index and E/E’ ratio and E/A ratio. r: Pearson correlation coefficient. NAMA, normal-attenuation muscle area; TAMA, total abdominal muscle area.

**Table 3.** Association between NAMA/TAMA index and LVEF, LV mass index, E/A ratio, and E/E’ ratio.

|  | Men |  |  | Women |  |  |
| --- | --- | --- | --- | --- | --- | --- |
|  | B | 95% CI | *p*-value | B | 95% CI | *p*-value |
| **LVEF** |  |  |  |  |  |  |
| Unadjusted | −0.004 | −0.019–0.010 | 0.564 | −0.018 | −0.033–−0.003 | 0.017 |
| Model 1<sup>a</sup> | −0.002 | −0.016–0.015 | 0.955 | −0.011 | −0.027–0.006 | 0.212 |
| Model 2<sup>b</sup> | 0.001 | −0.015–0.016 | 0.939 | −0.010 | −0.027–0.007 | 0.239 |
| Model 3<sup>c</sup> | 0.004 | −0.013–0.021 | 0.651 | 0.001 | −0.021–0.021 | 0.994 |
| **LV mass index** |  |  |  |  |  |  |
| Unadjusted | −0.069 | −0.135–−0.004 | 0.038 | −0.304 | −0.368–−0.240 | <0.001 |
| Model 1<sup>a</sup> | −0.015 | −0.082–0.053 | 0.668 | −0.111 | −0.181–−0.041 | 0.002 |
| Model 2<sup>b</sup> | 0.026 | −0.041–0.094 | 0.445 | −0.091 | −0.161–−0.022 | 0.010 |
| Model 3<sup>c</sup> | 0.041 | −0.035–0.117 | 0.289 | −0.053 | −0.139–−0.033 | 0.224 |
| **E/A ratio** |  |  |  |  |  |  |
| Unadjusted | 0.010 | 0.008–0.011 | <0.001 | 0.014 | 0.012–0.015 | <0.001 |
| Model 1<sup>a</sup> | 0.005 | 0.004–0.006 | <0.001 | 0.005 | 0.004–0.006 | <0.001 |
| Model 2<sup>b</sup> | 0.004 | 0.003–0.005 | <0.001 | 0.005 | 0.003–0.006 | <0.001 |
| Model 3<sup>c</sup> | 0.003 | 0.001–0.004 | <0.001 | 0.002 | 0.001–0.004 | 0.001 |
| **E/E' ratio** |  |  |  |  |  |  |
| Unadjusted | −0.047 | −0.055–−0.038 | <0.001 | −0.080 | −0.090–−0.070 | <0.001 |
| Model 1<sup>a</sup> | −0.026 | −0.035–−0.018 | <0.001 | −0.037 | −0.047–−0.026 | <0.001 |
| Model 2<sup>b</sup> | −0.019 | −0.027–−0.010 | <0.001 | −0.032 | −0.043–−0.022 | <0.001 |
| Model 3<sup>c</sup> | −0.012 | −0.022–−0.002 | 0.015 | −0.026 | −0.039–−0.013 | <0.001 |

<sup>a</sup>Model 1 adjusted for age.<sup>b</sup>Model 2 adjusted for variables in model 1 plus drinking, smoking, exercise habits, hypertension, and diabetes mellitus.<sup>c</sup>Model 3 adjusted for variables in model 2 plus GGT, triglyceride, LDL-C, hsCRP, and VSR.**Abbreviations:** LVEF, left ventricular ejection fraction; GGT, gamma-glutamyltransferase; LDL-C, low-density lipoprotein-cholesterol; hsCRP, high-sensitivity C-reactive protein; VSR, visceral to subcutaneous fat ratio.

## 4. Discussion

In this study, we analyzed the association between myosteatosis and cardiac function in an asymptomatic Korean population using a large cohort from health examination data. Individuals with lower NAMA/TAMA index values exhibited significantly lower E wave and E/A ratio and higher LA diameter, A wave, and E/E’ ratio, suggesting worse diastolic function. Multiple regression analysis confirmed that the NAMA/TAMA index was positively associated with the E/A ratio and negatively associated with the E/E’ ratio, even after adjusting for various covariates. To the best of our knowledge, this study is the first to analyze the impact of myosteatosis on cardiac function.

Individuals with a low NAMA/TAMA index, indicative of greater myosteatosis and poorer muscle quality, exhibited poorer LV diastolic function without significant changes in LV mass index and systolic function. These results support the hypothesis that myosteatosis may contribute to diastolic dysfunction through mechanisms similar to those observed in diabetic cardiomyopathy. ^8^ Diabetic cardiomyopathy initially manifests as educed early diastolic filling and increased atrial filling, due to exacerbated cardiac stiffness and impaired LV relaxation. ^19^ At this early stage, substantial changes in myocardial structure, such as LV mass and wall thickness, may not yet occur, ^20^ and LVEF is generally preserved. ^8,21,22^ This pathophysiological process could explain the lack of significant association between myosteatosis and both LV mass index and LVEF in the current analysis.

Among the covariates, age, sex, hypertension, diabetes mellitus, triglyceride, and VSR were independently associated with the E/E’ ratio (Supplementary Table S1). These variables have been shown to significantly impact diastolic function in previous studies, aligning with our findings. ^23–26^ In our study, the association between myosteatosis and diastolic function remained significant even after adjusting for these variables. These findings indicate that myosteatosis might affect cardiac function through mechanisms distinct from these traditional risk factors.

The underlying mechanism connecting myosteatosis and cardiac dysfunction is thought to be multifactorial. One potential mechanism is that systemic insulin resistance, caused by myosteatosis, leads to metabolic derangement and functional impairment of cardiomyocytes. When fat accumulates in the myocytes of skeletal muscle, it impairs insulin signaling and glucose uptake, contributing to systemic insulin resistance. ^27,28^ This insulin-resistant state can induce cardiac dysfunction without other underlying heart diseases, and even without overt diabetes mellitus. ^8^ Insulin resistance triggers a set of metabolic disturbances in cardiomyocytes, including impaired free fatty acid metabolism, stimulation of renin-angiotensin-aldosterone system, oxidative stress, and impaired calcium homeostasis. ^20,29^ These disruptions ultimately lead to the maladaptation of cardiomyocytes, resulting in cardiac hypertrophy and fibrosis, cardiomyocyte death, and eventually heart failure. ^20^ However, we were unable to confirm this pathophysiological mechanism linking myosteatosis-induced insulin resistance to cardiac dysfunction.

Another probable explanation for the association between myosteatosis and cardiac dysfunction is co-existing cardiac steatosis, in which fat deposition within the heart muscle itself impairs its function independently of systemic insulin resistance. Ectopic fat deposits in the heart may manifest as pericardial, either epicardial or peri-coronary, or myocardial steatosis. ^30^ Unlike myosteatosis that predominantly affects systemic metabolism, peri- or intra-myocardial lipid accumulation exerts primarily local adverse effects on the heart. ^31^ Myocardial triglyceride accumulation could lead to myocardial apoptosis and ventricular systolic dysfunction, ^31,32^ while an increase in epicardial adipose tissue also leads to ventricular hypertrophy, apoptosis, and impaired diastolic function. ^33,34^ Cardiac steatosis has been proposed as an independent predictor of diastolic dysfunction irrespective of BMI or the amount of visceral fat. ^35^ However, it is still challenging to determine whether cardiac steatosis is a fundamental cause of cardiac dysfunction or merely a consequence of metabolic derangement. ^30^ Although we could not directly assess the degree of cardiac steatosis in this study, we propose that ectopic fat in skeletal muscle might be a surrogate marker for fat accumulation in the heart, leading to similar detrimental effects. Further research is required to better understand the complex relationship between myosteatosis, cardiac steatosis, and cardiac function.

This study had several limitations. First, due to the retrospective and cross-sectional design of the study, we could not identify a causal relationship between myosteatosis and cardiac dysfunction. Second, we were unable to evaluate the degree of cardiac fat, which might be crucial for elucidating the underlying pathophysiological mechanisms connecting myosteatosis and cardiac function. Third, while CT imaging is considered accurate for assessing myosteatosis, we could not compare the degree of myosteatosis with other diagnostic modalities such as magnetic resonance spectroscopy. Lastly, the study population comprised relatively healthy, asymptomatic individuals undergoing routine health check-ups, which may restrict the generalization of our results to other populations or those with existing health conditions. Despite these limitations, our study has strengths in that it offers novel insights into the impact of myosteatosis on cardiac function and reveals the specific pattern of cardiac dysfunction associated with myosteatosis.

## 5. Conclusions

In this study, we demonstrated that the degree of myosteatosis was significantly associated with diastolic function in an asymptomatic population, while systolic function remained unaffected. These findings suggest that ectopic fat deposition in skeletal muscle may be an important predisposing factor in cardiac dysfunction. Future research is warranted to validate the pathophysiological mechanisms underlying the link between myosteatosis and diastolic dysfunction in more detail. Additionally, it would be valuable to investigate the impact of therapeutic interventions targeting myosteatosis on cardiac function. Understanding these mechanisms and potential treatments could have substantial implications for preventing and managing cardiac dysfunction associated with metabolic disorders.

## Data Availability

The data that support the findings of this study are available from the corresponding author, C.H.J., upon reasonable request.

## Acknowledgment

None

